# Policy implementation gap: a qualitative evaluation of the implementation of Ghana’s national cancer control strategy

**DOI:** 10.64898/2026.08.28.26361597

**Authors:** J. Bashiru, L. Baatiema

**Affiliations:** Department of Health Policy, Planning and Management, School of Public Health, University of Ghana, Legon, Accra, Ghana

**Keywords:** Cancer control, health policy, implementation, Ghana, National Cancer Control Strategy

## Abstract

**Background:** Cancer is a growing public health challenge in Ghana, with 27,385 new cases and 17,944 deaths recorded in 2022. Ghana developed a National Cancer Control Strategy (NCCS) in 2011 to guide prevention, early detection, treatment, and palliative care. The strategy expired in 2016 and has not been formally evaluated or renewed, leaving cancer control efforts without a guiding policy framework for nearly a decade. This study examined how the strategy was implemented, what barriers were encountered and what stakeholders recommend for a strengthened national cancer response.

**Methods:** We conducted a qualitative descriptive study using semi-structured key informant interviews. Fifteen participants were recruited through purposive sampling, supplemented by snowball referrals, representing three groups: Ministry of Health policymakers, frontline healthcare providers and representatives of cancer-focused non-governmental organisations. Data were collected between June and September 2025 and analysed using Braun and Clarke’s six-phase thematic analysis framework, guided deductively by the WHO Health Systems Building Blocks framework

**Results:** Three themes emerged: NCCS interventions and systems implemented, capturing progress in cancer awareness, HPV vaccination and pilot screening programmes alongside persistent geographic and financial inequities in access; barriers to implementation, including inadequate financing, infrastructure and workforce shortages, the absence of a national cancer registry and governance failures, among them the finding that no frontline healthcare provider interviewed had any awareness of the NCCS; and recommended implementation strategies, including co-production of a renewed strategy, establishment of a dedicated National Cancer Control Programme, expanded health insurance coverage and decentralisation of oncology services.

**Conclusion:** The NCCS was not operationally embedded in the health system. The evidence points to failures in policy dissemination as a constraint that precedes resource constraints. Addressing Ghana’s rising cancer burden requires renewed political commitment, co-produced governance structures and accountability mechanisms. These findings have relevance for other low- and middle-income country settings facing similar challenges.

## Introduction

Non-communicable diseases are now a leading cause of death in sub-Saharan Africa, and cancer is among the fastest-growing contributors to this burden. Globally, approximately 20 million new cancer cases and 9.7 million deaths were recorded in 2022 [1]. Low- and middle-income countries accounted for more than 70% of cancer-related deaths in 2020, driven by late-stage diagnosis, inadequate treatment infrastructure, and the absence of effective prevention systems [2]. In Ghana, 27,385 new cancer cases and 17,944 deaths were documented in 2022, reflecting a steady increase in both incidence and mortality over the preceding decade [1]. The annual economic loss attributable to cancer was estimated at USD 1.26 billion in 2019 [3]. Individual treatment costs are staggering relative to household incomes. One study found that breast cancer patients spent a median of GHC 31,021 within a single year of treatment, at a time when the national minimum daily wage was GHC 12.53 [4].

At the global and regional levels, several concerted efforts have been undertaken to strengthen the response to the growing cancer burden. The World Health Organization’s Global Action Plan for the Prevention and Control of Noncommunicable Diseases (2013–2030) [5] and the Global Initiative for Cancer Registry Development have guided countries in developing evidence-based national cancer control programs that emphasize prevention, early detection, and equitable access to treatment. In sub-Saharan Africa, initiatives such as the African Cancer Coalition, in collaboration with the National Comprehensive Cancer Network (NCCN) and the American Cancer Society, have supported the adaptation of cancer treatment guidelines to regional contexts. Additionally, several countries including Rwanda and Kenya have made progress by integrating HPV vaccination, cervical screening, and chemotherapy services into broader health systems. Despite these regional advances, implementation gaps persist due to inadequate financing, limited diagnostic capacity, and weak data systems challenges that Ghana similarly faces in its own cancer control effort

To address the growing cancer burden, Ghana’s National Cancer Control Strategy (NCCS) was developed in 2011 and formally launched in 2012. It was designed to address this burden through a structured, WHO-aligned framework covering five domains: prevention, early detection, diagnosis, treatment, and palliative care [6]. It set targets across these five domains for the period 2012 to 2016. It expired at the end of that period, has not been renewed, and no formal evaluation of its implementation has been published.

This is a significant problem. The implementation science literature is clear that even technically sound health policies regularly fail in practice when they are not supported by adequate health system capacity, governance structures, and contextual adaptation [7].

Existing Ghanaian cancer research has produced useful epidemiological assessments and policy critiques [8,9], but has not systematically examined how the NCCS has been operationalised, what obstacles have been encountered, or what those closest to implementation believe must change. This study addresses those questions.

Drawing on stakeholder perspectives from policymakers, healthcare providers, and civil society organisations, this study aimed to: (1) identify interventions implemented under the NCCS; (2) explore health system and governance barriers hindering implementation; and (3) capture recommended strategies for strengthening the national cancer response.

## Materials and methods

### Study design

A qualitative descriptive design was employed. This design was appropriate for the study’s aims: it enables in-depth, contextualised exploration of implementation processes, stakeholder experiences, and barriers to policy uptake that cannot be meaningfully captured through quantitative instruments. Qualitative methods are particularly well-suited to implementation research in complex health system contexts.

### Patient and public involvement

Patients and the public were not involved in the design or conduct of this study. Participants were policymakers, healthcare providers and civil society representatives, selected for their direct knowledge of cancer control implementation. Incorporating patient and survivor perspectives is a priority for future work.

### Selection and recruitment of participants

We used purposive sampling to identify 15 participants with direct experience of NCCS formulation, implementation or oversight, representing three groups: four Ministry of Health policymakers, six frontline healthcare providers and five NGO representatives. Initial contacts at the Ministry of Health were established through unit heads. Clinical participants were identified through oncology departments at study facilities, with initial contact made through department heads or unit coordinators. NGO participants were identified through established cancer-focused civil society networks, with snowball referrals used to reach additional participants.

Eligibility required a minimum of two years of direct engagement with cancer policy, service delivery or civil society advocacy in Ghana. Potential participants were contacted by both authors via telephone or institutional email and provided with a participant information sheet prior to consent. Written informed consent was obtained from each participant before their interview. All 15 individuals approached agreed to participate. This multi-level composition was designed to enable triangulation of perspectives across the policy, service delivery, and civil society dimensions of NCCS implementation. Professional experience ranged from 3 to 20 years. Participant characteristics are summarised in Table 1. Thematic saturation was assessed throughout data collection and was considered reached at 15 participants, as no substantively new themes or perspectives emerged in the final interviews.

**Table 1.**
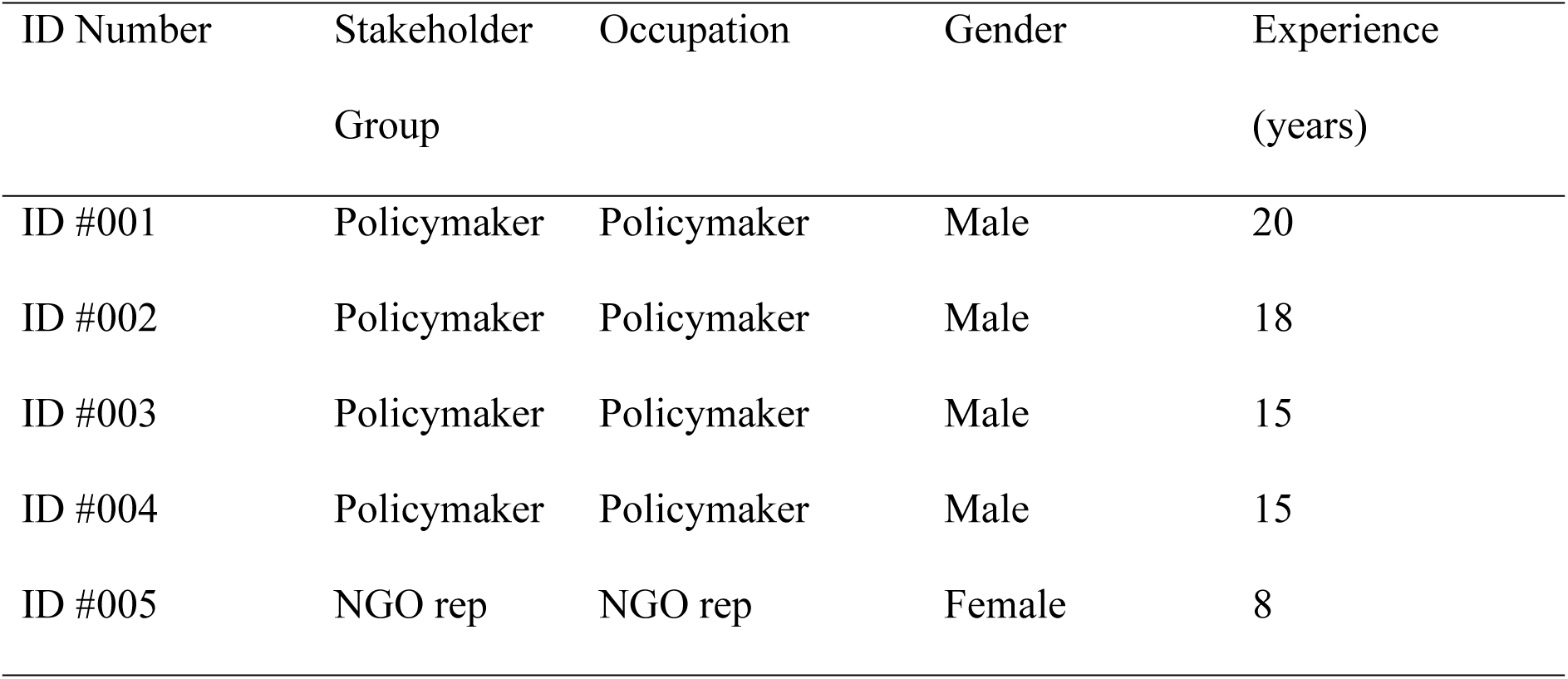
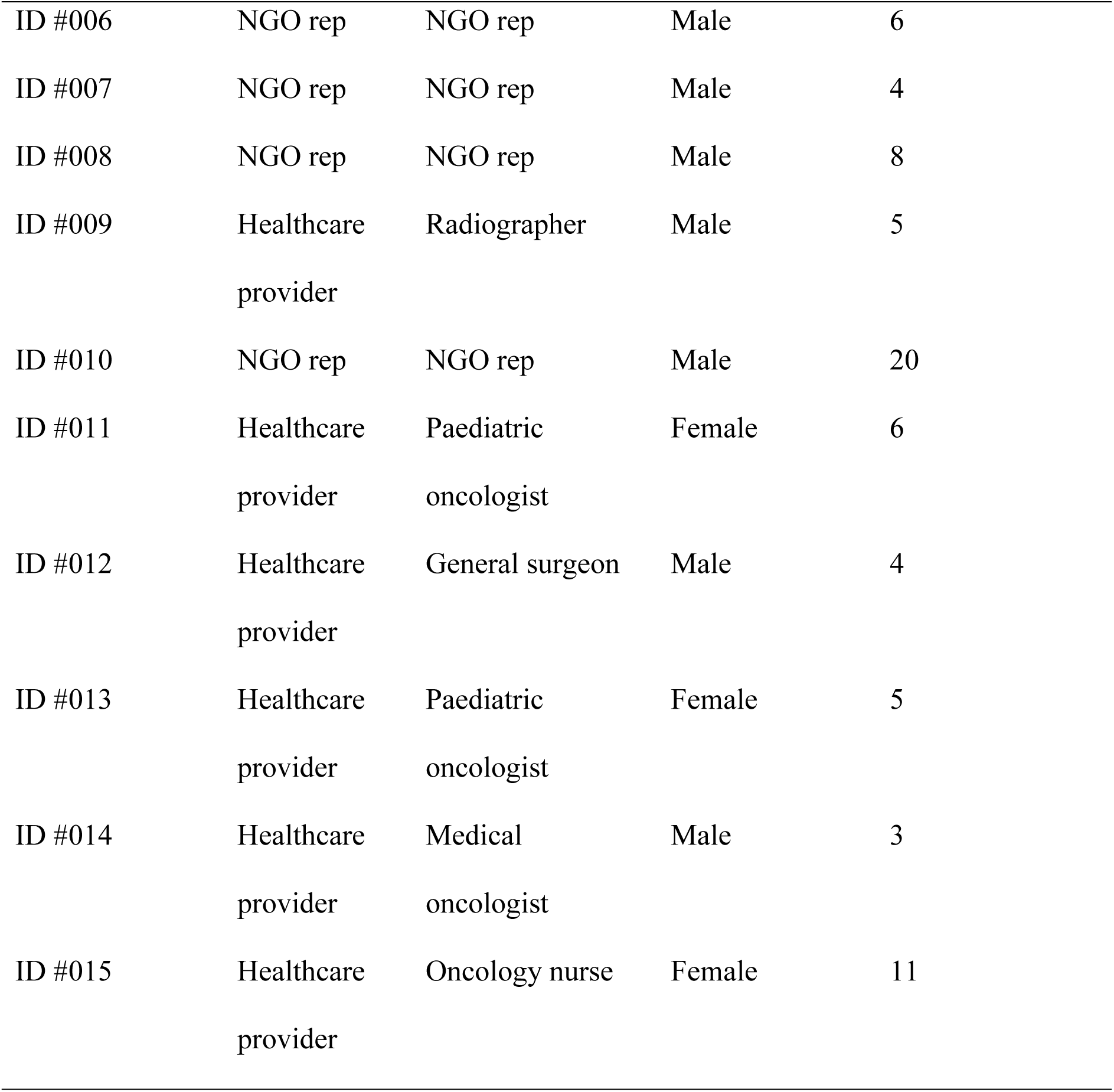
Characteristics of study participants (n=15)

| ID Number | Stakeholder Group | Occupation | Gender | Experience (years) |
| --- | --- | --- | --- | --- |
| ID #001 | Policymaker | Policymaker | Male | 20 |
| ID #002 | Policymaker | Policymaker | Male | 18 |
| ID #003 | Policymaker | Policymaker | Male | 15 |
| ID #004 | Policymaker | Policymaker | Male | 15 |
| ID #005 | NGO rep | NGO rep | Female | 8 |
| ID #006 | NGO rep | NGO rep | Male | 6 |
| ID #007 | NGO rep | NGO rep | Male | 4 |
| ID #008 | NGO rep | NGO rep | Male | 8 |
| ID #009 | Healthcare<br>provider | Radiographer | Male | 5 |
| ID #010 | NGO rep | NGO rep | Male | 20 |
| ID #011 | Healthcare<br>provider | Paediatric<br>oncologist | Female | 6 |
| ID #012 | Healthcare<br>provider | General surgeon | Male | 4 |
| ID #013 | Healthcare<br>provider | Paediatric<br>oncologist | Female | 5 |
| ID #014 | Healthcare<br>provider | Medical<br>oncologist | Male | 3 |
| ID #015 | Healthcare<br>provider | Oncology nurse | Female | 11 |

### Study setting

The study was conducted across participants drawn from tertiary healthcare facilities with oncology services: Korle Bu Teaching Hospital and Greater Accra Regional Hospital in Accra; Cape Coast Teaching Hospital in Cape Coast; Tamale Teaching Hospital in Tamale; and the Sweden Ghana Medical Center (private) in Accra. The Ministry of Health was included as the primary policy institution. Four civil society and NGO partners were included: World Child Cancer, the Clinton Health Access Initiative, Ghana NCD Alliance, and City Cancer Challenge.

### Study tool

Three semi-structured interview guides were developed following an extensive literature review and consultations with Ministry of Health officials, with separate versions for policymakers, healthcare providers, and NGO representatives, reflecting their differing vantage points on NCCS implementation. Guides were developed to explore: the nature and extent of NCCS implementation activities; perceived barriers to implementation across health system domains; and stakeholder recommendations for strengthening the national cancer response. Guides were pilot-tested with two individuals not included in the final sample.

### Data collection

All interviews were conducted in English by the first author (JB), under the supervision of the second author (LB), who has extensive experience in qualitative research. between June and September 2025, lasting 50 to 120 minutes. Guides were pilot-tested with two individuals not included in the final sample. Interviews took place at participants’ workplaces or virtually where face-to-face contact was not feasible. All interviews were audio-recorded with informed consent, transcribed verbatim, and cross-checked against recordings for accuracy. All participants provided written informed consent before taking part. Participants were aware of the interviewer’s professional role and research aims, and verbal rapport was established during initial contact and maintained throughout each interview. No repeat interviews were conducted. Field notes were made during and immediately after each interview to capture contextual observations relevant to the analysis. This study followed the Consolidated Criteria for Reporting Qualitative Research checklist.

### Data analysis

Thematic analysis was conducted using the six-phase framework of Braun and Clarke [10]. In Phase 1 (familiarisation with data), JB read and re-read all 15 transcripts in full, making initial reflective notes and identifying preliminary patterns of interest. In Phase 2 (generating initial codes), a systematic line-by-line coding process was applied to the entire dataset, producing a comprehensive set of initial codes capturing both the semantic content and interpretive meaning of participants’ accounts. In Phase 3 (searching for themes), codes were collated and grouped into candidate themes, and a thematic map was developed to explore relationships between emerging categories. In Phase 4 (reviewing themes), candidate themes were reviewed against the coded data extracts and the full dataset to assess their internal coherence, distinctiveness, and grounding in participant accounts; themes that did not hold up across the dataset were refined or merged. In Phase 5 (defining and naming themes), a clear analytic narrative was developed for each theme, with names chosen to reflect both the content and interpretive scope of the theme. In Phase 6 (producing the report), the final themes were written up with illustrative participant quotations selected to represent the breadth and range of perspectives within each theme was both deductive, drawing on the WHO Health Systems Building Blocks framework [11] as an organising lens, and inductive, allowing unanticipated themes to emerge from participants’ accounts. NVivo 14 facilitated systematic data management.

### Rigor and trustworthiness

The trustworthiness of the study was maintained, in accordance with Guba and Lincoln’s proposition in 1985 by ensuring that the study was credible, transferrable, dependable, and confirmable [12]. Member checking (participants reviewed summaries) of the study, prolonged engagement with transcripts and the field, and triangulation with field notes was done to enhance credibility. For purposes of transferability, rich context description was carried out. Dependability and confirmability were ensured through audit trail of coding decisions, peer debriefing, and reflexive memoing

### Ethical approval

Ethical clearance was obtained from the Ghana Health Service Ethics Review Committee (GHS-ERC: 028/03/25). Informed consent was obtained in writing from each participant prior to the interview. Data are anonymised; participants are identified only by a unique alphanumeric code and broad occupational category. Audio recordings and transcripts were stored securely in accordance with institutional data protection protocols. The study was conducted in accordance with the ethical principles of the Declaration of Helsinki.

## Results

### Characteristics of the participants

Fifteen participants were interviewed across three stakeholder groups. Four were Ministry of Health policymakers with direct involvement in NCD and cancer policy development. Six were frontline healthcare providers (two paediatric oncologists, one medical oncologist, one general surgeon, one radiographer, and one oncology nurse). Five were representatives of civil society and NGO organisations. Eleven participants were male. Professional experience ranged from 3 to 20 years. All interviews were conducted in English. Thematic saturation was considered reached at 15 participants, with no substantively new themes emerging in the final two interviews. Table 1 presents participant characteristics.

### Overview of the results

Three major themes emerged from the data describing how NCCS implementation has unfolded in Ghana since 2012. These included: (1) NCCS interventions implemented; (2) barriers to implementation, encompassing health system and governance dimensions; and (3) recommended implementation strategies. Each theme is presented below, supported by illustrative participant quotations. Themes and subthemes are summarised in Table 2.

**Table 2.**
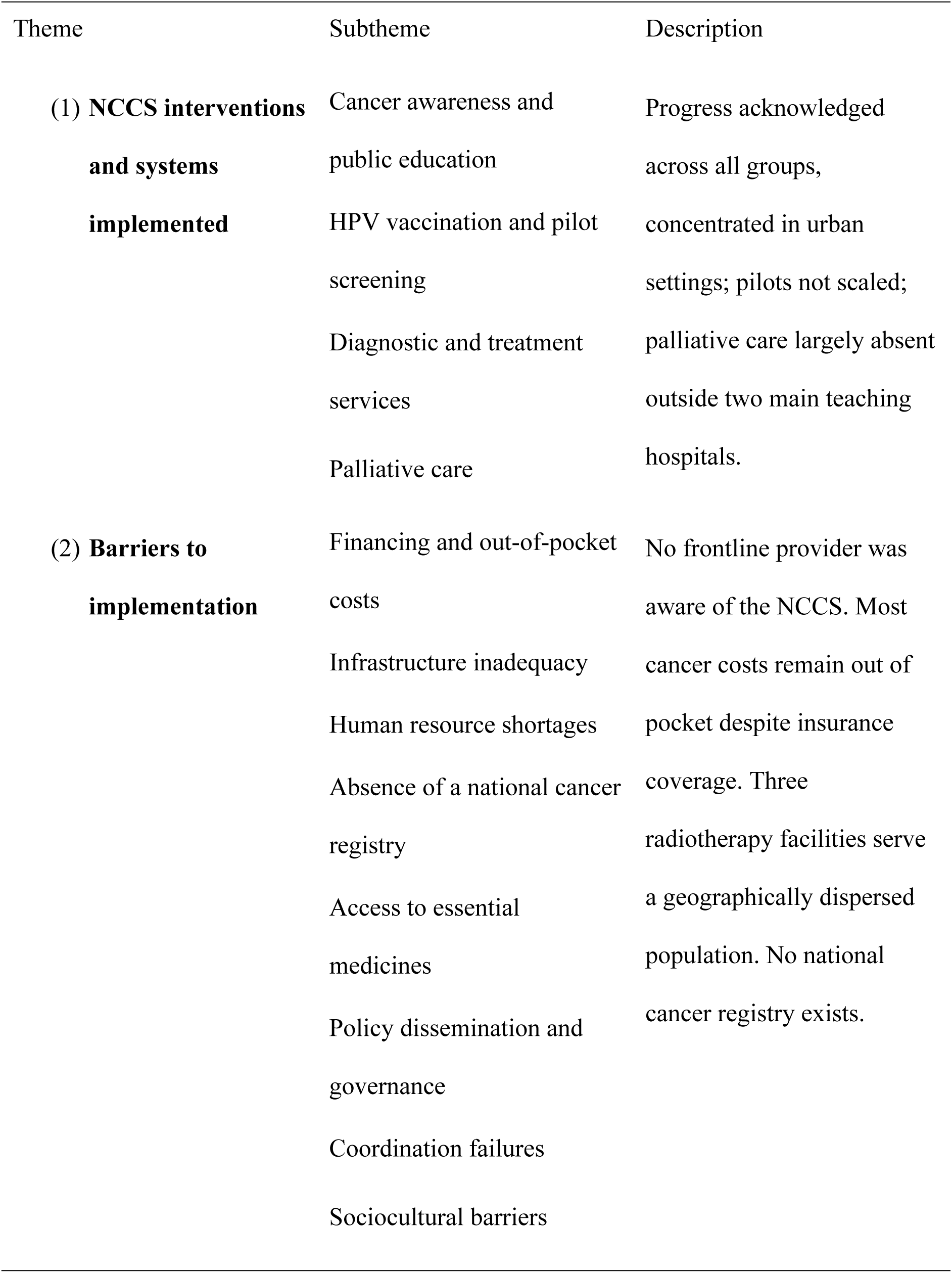
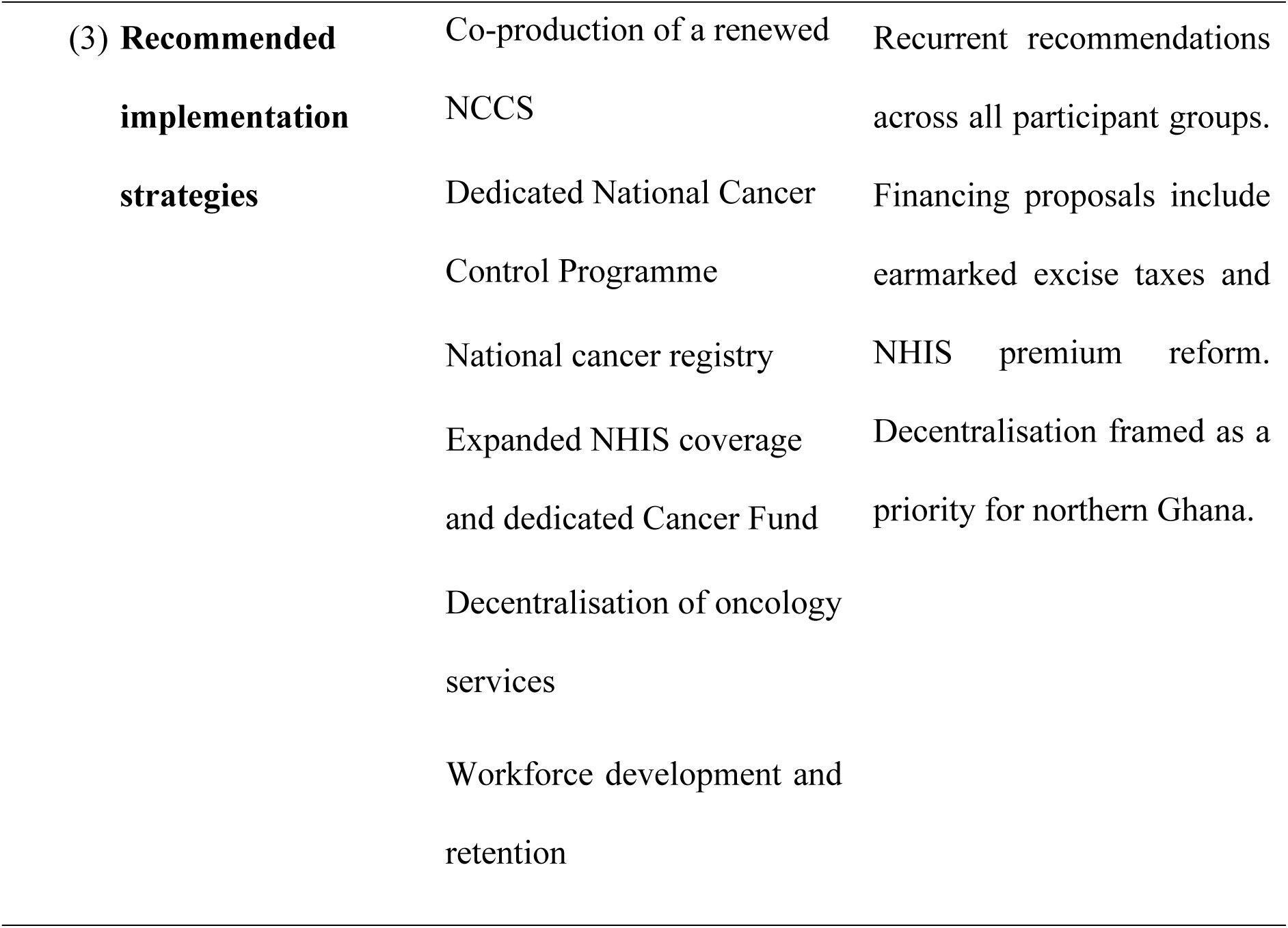
Themes and subthemes.

### Theme 1: NCCS interventions and systems implemented

#### Sub-theme 1: Cancer awareness and public education

Participants across all groups acknowledged measurable progress in public cancer awareness since the NCCS’s launch. Annual observances such as Breast Cancer Awareness Month, expanded radio and television coverage, and targeted community campaigns were cited as evidence of change. This progress was consistently qualified, with participants noting that campaigns have remained concentrated in urban centres and that rural populations have been largely underserved.

*Earlier, there was little information about these cancers. But now we know that there’s a lot of information that is on our radios, on our news, and we even have the Pink October, where we have the breast cancer awareness creation. So we are making progress in terms of awareness*. - **ID #008, NGO Rep**

#### Sub-theme 2: HPV vaccination and pilot screening initiatives

HPV vaccination and pilot cervical and breast cancer screening programmes were acknowledged, though access was described as limited. One policymaker described a pilot in which community health post nurses were trained to offer breast examinations as part of routine consultations, framing integration into primary care as a scalable approach. The pilot had not been scaled at the time of the study.

*Yesterday when I went to the field, I saw how the nurses there, it was a CHPS compound, Atankreja CHPS compound, where they have been trained to offer, as part of the project, which is going to be scaled up, they are trained to offer breast cancer awareness creation and screening for every mother who comes in contact with their facility. So it’s like you are integrating breast health care*… -**ID#004, Policymaker**

*Many women want to get screened, but the cost and distance discourage them. For those in the remote areas, screening centres are too far away, and even when they get there, mammograms are expensive. Unless it is subsidized or part of an outreach program, most people simply opt out* -**ID #005, NGO Rep**

#### Sub-theme 3: Diagnostic and treatment services

Diagnostic and treatment services were described as available but geographically concentrated at teaching hospitals in Accra and Kumasi. Referral pathways beyond those facilities were poorly defined. Providers reported pathology delays of up to two weeks, attributed to the absence of frozen section capability at most facilities.

*…we are unable to do frozen sections… So we have to wait, get the pathology report maybe two weeks later. There are some delays in getting some reports and that contributes to delay in starting treatment.* -**ID #012, General surgeon**

#### Sub-theme 4: Palliative care

Palliative care featured in the NCCS’s objectives but was universally described as deprioritised and practically absent. No trained palliative care specialist was identified at any facility outside the two main teaching hospitals.

*… We don’t have a trained palliative care specialist doctor. Neither do we have palliative care nurses…* -**ID #012, General surgeon**

### Theme 2: Barriers to implementation

#### Sub-theme 1: Financing and out-of-pocket costs

Financing was the most consistently and urgently cited barrier across all participant groups. Despite National Health Insurance Scheme (NHIS) coverage for breast and cervical cancers, and an announced extension to four childhood cancers in 2022, the majority of cancer care costs were reported to remain out of pocket. Surgery, radiotherapy, diagnostics and most chemotherapy drugs fall outside NHIS coverage. Participants described patients selling land, borrowing from family members and exhausting household savings to fund treatment.

*Breast cancer care is quite expensive… the average Ghanaian won’t be able to afford. Even though some chemotherapy agents are covered under the NHIS, patients still pay for diagnostics, surgery, and radiotherapy out of pocket, and these costs are catastrophic for many.* -**ID #012, General surgeon**

*It breaks my heart to see patients walk away from treatment, not because the cancer is incurable, but because the cost is. Families sell their homes, exhaust their savings, and still can’t afford to keep their loved ones alive. Cancer shouldn’t be a death sentence simply because one is poor.* -**ID #015, Oncology nurse**

#### Sub-theme 2: Infrastructure inadequacy

Infrastructure inadequacy was described consistently and in concrete terms. Ghana has only three radiotherapy facilities, all located in the south of the country. Patients from the Northern, Upper East, and Upper West regions must travel to access radiation therapy, and many do not complete treatment as a result.

*…Access to the services is an issue, because it is not readily distributed. If you live in a region where the service is only available at the regional capital, or in a region like Western where there is no teaching hospital, you need to travel… to access that service*. -**ID #003, Policymaker**

#### Sub-theme 3: Human resource shortages

Human resource shortages were described as acute across facility types. Oncology units reported staffing ratios that required providers to assume multiple roles simultaneously, with patient-to-nurse ratios described as unsafe.

*oh. Staffing, we are grossly understaffed. We need more people to come into oncology. We need more nurses to specialize in oncology…So I mean, under standard circumstances, we should have one nurse, one patient because most of them are really ill.* -**ID #011, Paediatric Oncologist**

#### Sub-theme 4: Absence of a national cancer registry

The absence of a national cancer registry was identified as a foundational gap by every participant group. Cancer incidence data for Ghana currently rely on modelled estimates drawn from data in neighbouring countries. Policymakers, providers, and NGOs all described the same consequence: decisions are made without reliable evidence.

*Without data, it’s hard to know which cancer is killing us most and where to intervene.* -**ID #002, Policymaker**

#### Sub-theme 5: Access to essential medicines

Shortages of chemotherapy drugs and the absence of domestic HPV vaccine production were raised across participant groups. Participants noted that reliance on imported vaccines creates supply vulnerabilities and that the affordability of cancer medicines remains a barrier for patients without insurance coverage.

*…access to quality and affordable medications or treatment is also a very big challenge.* -**ID #007, NGO Rep**

*“So, currently in Ghana, we do not produce… domestically, we do not produce vaccines for HPV. Like, we do not have that vaccine. So, domestic vaccine production is a challenge.* -**ID #003, Policymaker**

#### Sub-theme 6: Policy dissemination and governance failures

The single most striking finding in this study is the following: every frontline healthcare provider interviewed had no awareness of the NCCS whatsoever. Not one had heard of it, read it, or been briefed on it. This is not a coordination failure in the conventional sense. It means the strategy was formulated at the policy level and never reached the people who were expected to implement it. The NCCS expired in 2016 and has not been renewed. The absence of a current cancer-specific national framework was described by participants across all groups as producing a governance vacuum.

*In terms of my main work as a clinical person, nobody has spoken to us about it. We don’t know what is in it. We don’t know what it is.* **-ID #009, Radiographer**

*So, we don’t have any kind of updated version. What that means is that you don’t know whether you are to follow or work with the old strategy … Because that’s like we are almost getting to 10 years since it expired. There’s a broader NCD policy, but you don’t get a lot of specifics.* -**ID #007, NGO Rep**

#### Sub-theme 7: Coordination failures

Coordination structures envisioned in the NCCS were described as non-functional.NGO participants described being excluded from policy development processes and presented with finalised documents for endorsement rather than contributing to their design. The NHIS childhood cancer benefit announced in 2022 was reported by providers as still not operational at the time of data collection.

*There are instances with no consultation – they develop the policies and then say we should come and endorse… -***ID #010, NGO Rep**

*We are hearing there are some hitches here and there. Our mothers are not benefiting from it. They are still buying the chemotherapy medications. They are still buying them, expensive* -**ID #015, Oncology nurse**

#### Sub-theme 8: Sociocultural barriers

Participants described patients discontinuing medical treatment to pursue traditional or spiritual healing and returning to hospital only after disease had advanced. Community representatives raised concerns about short-term programme visits that did not result in sustained follow-through.

*So some of them, we start the treatment, they are doing well and they go and continue with their traditional healers. And then they try something. Even their pastors and spiritual people also try that. When they realise that it’s beyond them, they now tell them, no, at this point you have to go to the hospital.* -**ID #011, Paediatric oncologist**

*when you come, we don’t see you again…you come, take your data and go. People come every day and no change happens* -**ID #010, NGO Rep**

### Theme 3: Recommended implementation strategies

Three recommendations recurred with near unanimity across all participant groups. First, the urgent revision of the NCCS through a genuine co-production process that involves healthcare providers, NGOs, communities, and policymakers from the outset. Second, the establishment of a dedicated National Cancer Control Programme with a named programme manager, a defined mandate, and regular multi-stakeholder coordination. Third, the establishment of a national cancer registry as the foundation for evidence-based policy and outcome monitoring.

*…get everybody on board when you are developing a document. Not only at the last stage… that’s called co-production, Get everybody involved and then begin the process.* **-ID #010, NGO Rep**

*We need to have a cancer control programme. Either it sits at the ministry, or it sits at the Ghana health service. We need a dedicated cancer control programme, which would coordinate all the cancer control activities* -**ID #007, NGO Rep**

*“maybe we may have to revisit the strategy of creating a national cancer registry. Because I think that if we have something like that in place, it will be able to bring out the core truth or the core facts. Because the numbers will let us know nationwide what the real issues are.* -**ID #003, Policymaker**

On financing, specific proposals included expanding NHIS coverage to prostate, colorectal, and liver cancers; subsidising screening and preventive services; and raising the NHIS premium above its current GHC 23 annual rate. Participants also called for a dedicated Cancer Fund financed in part by earmarked excise taxes on tobacco, alcohol, and sugar-sweetened beverages.

*23 cedis a year is not realistic. It cannot take care of treatment if you have a cancer. And the cost of these medications….it is not realistic… even for malaria treatment. **-*****ID #004, Policymaker**

*I mean the excise taxes that the government have put on tobacco and alcohol and SSBs. The government should earmark 50% of that money to support the Ghana Medical Trust Fund.* -**ID #007, NGO Rep**

Service decentralisation was a consistent theme across all groups, with stakeholders calling for regional oncology centres with full radiotherapy and chemotherapy capacity, and particular urgency expressed for northern Ghana. Workforce recommendations focused on training more specialists, equitable geographic posting, and retention incentives to address brain drain. For prevention and early detection, free or subsidised screening for all common cancers, full institutionalisation of HPV vaccination, and community engagement strategies involving traditional leaders, religious institutions, and schools were identified as essential.

## Discussion

This study provides one of the first systematic qualitative evaluations of NCCS implementation in Ghana. The findings reveal that more than a decade after the strategy’s launch, its ambitions have not been operationalised, its frontline workforce was never informed of it, and the governance architecture required to implement it was never built. Three themes characterise the implementation landscape: measurable but geographically limited progress in awareness, vaccination, and pilot screening; severe and compounding barriers across financing, infrastructure, human resources, data systems, governance, and sociocultural alignment; and a clear consensus among stakeholders on what a reformed national cancer response must entail.

The complete absence of NCCS awareness among frontline providers is the most consequential finding. Prior critiques have noted that the strategy lacked evaluation and had not been updated [8]. What this study adds is more fundamental: the strategy was never communicated to those expected to implement it. The policy-to-practice gap in Ghana’s cancer response begins much earlier than resource constraints, at the point where a strategy is formulated at the centre and never reaches the frontline. This finding does not appear in the existing literature and has direct implications for how Ghana designs and disseminates its next strategy.

The prevention and screening findings are consistent with published evidence. Ninety-seven percent of Ghanaian women have never had cervical cancer screening [13]. Rwanda, a country with comparable resource constraints, achieved over 90% HPV vaccination coverage through a school-based programme built on sustained political commitment at the highest level and structured private sector engagement [14]. The contrast illuminates what differentiates policy intent from policy delivery. Ghana had comparable commitments in the NCCS. What was absent was the institutional architecture to translate them into a sustained national campaign. The infrastructure and workforce findings align with documented regional evidence. Geographic accessibility research confirmed that fewer than 47% of Ghanaians live within 100 km of a radiotherapy centre, and an estimated 23 additional machines are needed nationally [15]. The health system carried a 41% workforce vacancy rate in 2018 [16]. Fewer than ten radiation oncologists served the public sector in the preceding decade. These are not new findings. What this study demonstrates is their persistence despite a decade of strategy commitments. The constraint is not knowledge of the gap. It is the absence of the political mechanism to close it.

A critical barrier identified in this study is the absence of a comprehensive national cancer registry, which continues to undermine effective cancer control efforts in Ghana. The lack of reliable, population-based data means that current estimates of cancer incidence are largely modelled or extrapolated from neighbouring contexts, limiting the ability of policymakers and healthcare providers to make evidence-based decisions regarding planning, resource allocation, and service delivery. This gap also raises broader governance concerns, as weak data systems increase the risk of policy priorities being influenced by external actors rather than grounded in the country’s epidemiological realities. Although the NCCS acknowledged the establishment of a population-based cancer registry as a key priority, progress has remained slow. This finding aligns with existing literature, which identifies inadequate cancer data systems as a persistent constraint to effective cancer planning and monitoring in low- and middle-income countries [17]. Overall, the absence of a functional national cancer registry represents not merely a technical gap, but a fundamental weakness in the governance, monitoring, and accountability mechanisms of cancer control in Ghana.

The sociocultural findings deserve more than footnote treatment in cancer policy. Systematic review evidence documents that many Ghanaian women interpret breast cancer through spiritual frameworks before seeking hospital care [18]. An estimated 80% of the sub-Saharan African population uses traditional medicine [19]. These are not peripheral anomalies. They are central features of the care-seeking context in which cancer policy must operate. The current NCCS acknowledged cultural barriers. It did not design mechanisms to address them. A revised strategy must do both.

The financing proposals from stakeholders are consistent with available evidence. NHIS coverage currently falls short of population need and excludes many essential drugs and specialist procedures, driving catastrophic expenditure among cancer patients [20,21]. Earmarked health taxes and expanded benefit packages have been demonstrated as feasible mechanisms in comparable settings. The recommendation for a dedicated Cancer Fund reflects a pragmatic response to the constraint that expanding the NHIS alone is insufficient for the cost burden of cancer care.

The governance findings are also consistent with Baatiema et al’s analysis of NCD policy implementation in Ghana, which documented gaps between policy announcement and operational delivery across multiple disease areas [22]. The NHIS childhood cancer coverage finding in the current study is a precise and recent example of the same pattern. A benefit is announced. Families are told their children are covered. Three years later, they are still buying chemotherapy out of pocket. This suggests a governance failure that extends beyond financial constraints. Addressing it requires accountability mechanisms, not just additional funding.

The proposed strategies are consistent with international guidance. World Health Assembly resolutions on cancer explicitly call for national programmes with empowered programme managers, workforce development, and integrated data systems [23]. Countries with effective cancer control programmes have built exactly this institutional architecture. Ghana has called for it in successive policy documents. The call has not been answered. This study provides the qualitative evidence needed to make that failure visible and to inform the political argument for change.

For policy, the findings suggest that a renewed NCCS must be co-produced with the full range of implementing actors from the outset, accompanied by a named programme management structure and a dissemination process that treats frontline awareness as a core implementation task. For practice, structured orientation for all relevant healthcare cadres is needed alongside engagement of community health workers and primary care facilities in early detection and referral. For research, priorities include studies centred on patient and survivor perspectives, quantitative monitoring of implementation indicators and comparative analyses across sub-Saharan African settings facing similar cancer control challenges.

Several limitations should be considered. The study focused on tertiary facilities and national-level institutions; perspectives from district and community-level implementers in rural settings, where implementation gaps are likely most acute, were not captured and should be a priority in future work. The exclusion of cancer patients and survivors is a further limitation, as their perspectives would add a critical dimension to the implementation account. As with all qualitative designs, the findings are not statistically generalisable, though they are analytically transferable to settings with comparable health system characteristics. Social desirability bias, whereby participants may have moderated critiques of institutional processes, cannot be excluded.

## Conclusion

Ghana’s National Cancer Control Strategy was a sound and necessary policy. Its implementation was not sound. The strategy expired in 2016 without renewal, was not disseminated to frontline healthcare providers, and was not backed by the coordinating, financing, or infrastructure mechanisms needed to make it operational. The path forward is clear in the evidence: a renewed, co-produced, and properly resourced NCCS; a dedicated cancer control programme with institutional authority and real accountability; and the governance mechanisms to close the long-standing gap between policy intent and system performance. Ghana’s cancer burden will continue to grow. Whether it continues to go inadequately answered is a matter of political will.

## Data Availability

The de-identified datasets generated and analysed in this study are available from the corresponding author on reasonable request.

## Acknowledgements

The authors thank all study participants for their time and expertise. Thanks are also extended to the Ghana Health Service for facilitating access to study sites.

## Supporting information

**S1 Doc. Reflexivity statement**

**S2 Doc. Interview Guide/Questionnaire for Policymakers**

**S3 Doc. Health Providers (Oncologists, Nurses, Oncology Pharmacists) Interview Guide**

**S4 Doc. Interview Guide – NGOs/ Development Partners**

**S5 Doc. Consolidated criteria for reporting qualitative studies (COREQ): 32-item checklist**

